# Factors affecting oral health problems among school children in Kaski District, Nepal

**DOI:** 10.1101/2022.04.25.22274284

**Authors:** Kamal Prasad Chapain, Krishna Gopal Rampal, Kalpana Gaulee Pokhrel, Chiranjivi Adhikari, Deependra Hamal, Khem Narayan Pokhrel

## Abstract

**Introduction:** School children have a high prevalence of oral health problems in Nepal. Socio-demographic factors such as gender, parents’ socioeconomic status, and individual awareness of oral health may have an influence on the occurrence of oral health problems. However, little evidence is available about the oral health problems and their associated factors among school children. Therefore, this study aimed to assess the factors associated with oral health problems among school children in Nepal.

**Methodology:** A cross-sectional study was conducted among school children of grade 7 in 12 schools of Kaski district, Gandaki Province, Nepal. The schools were randomly chosen from urban and semi-urban areas in the district. A total of 669 students participated in the study. Data were collected using a set of questionnaires covering dental health knowledge, socio-demographic characteristics, oral health condition and practices. The factors of poor oral health condition and practices were examined using t-test, one-way Anova, and multiple linear regression.

**Results:** School children who visited health institutions for oral health services and those with parents having higher level education had higher dental health knowledge scores. Total decayed score was higher among those who did not have knowledge that fluoride prevents decay compared to those who had knowledge about it (Have knowledge about fluoride prevents decay: Mean=1.21 (SD=1.54) Vs No knowledge: mean=2.13 (SD=2.13). Females were more likely to have higher DMFT score compared to males (β-coefficient=0.43, 95% CI=0.13, 0.73, P value=0.005) and increase in knowledge score was associated a with a decrease in DMFT score (β-coefficient=-0.09, 95% CI= -0.20, -0.10, P value=0.047).

**Conclusion:** This study found that gender, knowledge score, and parent’s socioeconomic status were the major factors contributing to higher DMFT scores. School children should be provided with regular oral health counseling and promotion activities in schools and oral health screening in schools.

## Introduction

Oral health problems are common among school children. An estimated 60-90% of school children suffer from dental caries worldwide (WHO, 2016). Dental caries is the most prevalent condition included in the 2016 Global Burden of Disease Study, ranking first for the decay of permanent teeth (2.3 billion people) and 12th for deciduous teeth (560 million children). It has been found that children suffering from oral disease are 12 times more likely to be restricted in daily activities than normal children who don’t have any oral disease (1) Oral diseases are usually multifactorial. Socioeconomic factors affect the severity and the distribution of oral diseases. Family environment including socio-behavioural and environmental factors have a strong relationship with the oral health condition of children. Moghaddam et al., showed that children’s poor oral health condition is related to lower education level of their mother and father and with lower family income (2). Children of parents who have lower education level and lower level of socio-economic status are more affected by dental caries and other oral health related problems (3).

To promote oral health education, a supportive environment should be provided to the community as well as school children and policy makers should plan in such a way by which social discrimination can be reduced(4). Healthy oral heath behavior minimizes the risk of various oral health problems. Brushing twice a day with fluoride toothpaste is one of the most important habits for good oral health. Through brushing during the day and night after meal, children can learn about the benefits of good oral hygiene and are to be taught to brush their teeth twice daily with fluoride toothpaste (5).

Healthy oral health practice is one of the factors to prevent oral health problems. A study in Nepal shows almost 95 % school children reported cleaning their teeth at least once a day and 71% only used fluoridated tooth-paste. In this study it was found that during the last 6 months only 4% of them received service from oral health services (Thapa et al.,). Due to the low level of awareness, oral health education and oral hygiene practice, promotion for prevention is essential. Dental treatment is also highly expensive and difficult to be afforded by most of the people with low socioeconomic background. Although dental caries is a highly preventable disease, a high proportion of the people in Nepal who are poor and marginalized are suffering from oral health related problems. Poor oral hygiene is regarded as one of the most important risk factors for common oral diseases. The most common cause of deteriorating oral health conditions of Nepalese people are changes in dietary habit.

Many school children suffer from oral health problems in Nepal. A total of 58% of schoolchildren aged between 5-6 years were affected by dental caries according to report by the National Pathfinder Survey, 2004. Another study found that oral health related problems was the prime reason of missing school in Nepal where32% of the students missed their school due to dental pain (6).

Very few studies have examined the association between oral health and oral hygiene practices with sociodemographic factors. Although there are a few studies in Nepal which describe the socio-economic factors and oral health practices among school children that are contributing to oral health problems, the knowledge level of school students, practices in terms of using fluoridated toothpaste and knowledge about teeth and gums is not well known. Therefore, this study aimed to assess these factors associated with oral health problems among school children in Nepal.

## Methods

### Study design and setting

A cross sectional study was conducted among school children in Grade 7 in 12 community schools in the Kaski district of Nepal. Kaski district is the provincial capital of Gandaki Province in Western Nepal. The schools were located in urban and semi-urban areas of the district. We purposively selected Kaski district as the study district because Kaski district is the provincial headquarters, where people with diverse population from more than 15 districts of its catchment area live.

### Sampling strategy and sample size

We randomly selected 12 government schools out 325 schools. All children in Grade 7 of the selected schools were included in the study. We presumed the prevalence of dental caries is 60% among school children in calculating the estimated sample size considering 95% confidence and 80% power of the study. The estimated sample size was 654 and we increased the sample size to 669 in this study considering various characteristics to be studied.

### Participant’s inclusion and exclusion criteria

Those school children, who were enrolled in their school at least 6 months before the study in grade 7 were included. Those, who were not able to respond the questions because of their intellectual disability were excluded from the study.

### Study tools and measurements

#### Sociodemographic characteristics, oral health knowledge, and practices

Information on students’ sociodemographic characteristics, visiting dentist or dental care practitioners for their services, student’s knowledge on oral health, oral health practice and oral health seeking behavior were elicited using a set of questionnaires.

Student’s oral health knowledge was scored using a system adopted from previous studies (7,8) Oral health knowledge was determined by asking the following questions (1) periodontal disease can affect health, (2) regular tooth brush can protect tooth decay, (3) fizzy soft drinks affect the teeth, (4) use of fluorides prevent tooth decay, (5) gingivitis is a disease that makes your gums bleed, (6) causes of dental caries or proper tooth brushing can prevent dental caries (7) sugar causes tooth decay, (8) tooth decay is a disease that destroys your teeth, and (9) healthy teeth means strong and carries free teeth. Oral health practices were determined by asking the following questions: (1) frequency of brushing teeth, per day, (2) time spent for brushing in minute, (3) cleansing aid used, (4) materials used to clean teeth, (5) frequency of changing tooth brush, (6) type of toothpaste used, (7) mouth rinsing after eating, (8) clean tongue after meal or during rushing.

### Oral Health Condition (DMFT score)

Oral health condition of the student was assessed and expressed using the Total DMFT Index Score and Total Decayed Score. The DMFT score numerically expresses the prevalence of dental caries and is obtained by calculating the number of Decayed (D), Missing (M) and Filled (F) teeth (T). It is used to get an estimate how much the dentition until the day of examination has become affected. It is calculated for 28 (permanent) teeth, excluding third molar (wisdom teeth). Oral examination is carried out to determine: How many teeth have caries lesions (incipient caries not included), how many teeth have been extracted and how many teeth have fillings or crowns. A total score of DMFT is 28. (The Third edition of “Oral Health Surveys - Basic methods”, Geneva 1987).

### Data collection

Oral health condition was examined by the researcher and two research assistants collected the information from school children from April to May 2020. Prior to data collection the research assistants were given three days training covering data collection and ethical procedures. Pre-testing of questionnaires were done in non-selected schools and necessary modifications in questionnaires were made.

### Data analysis

Statistical analysis was conducted to examine the association between factors and dental health condition, knowledge, and practices of school children. Multivariate analysis was conducted to determine the factors associated with DMFT scores after controlling for sociodemographic variables. We used SPSS version 20 for the entire data analysis process.

### Ethical consideration

Ethical approval was obtained from the Nepal Health Research Council (ERB Protocol Registration No. 165/2021PhD) and the University of Cyberjaya (UOC) Research Ethics Committee (CRERC Reference no UOC/CRERC/EXTERNAL/07/2020 -SPC620200605). Signed consent from the participants and assent from the head teacher of the school were obtained. Participation was ensured to be completely voluntarily and confidential.

## Results

### Descriptive characteristics on the sociodemographic characteristics, oral health knowledge, oral health practice and oral health condition of school children

Of total participants (n=669), 54.9% were females, and 67.1% had used health services for oral health. More than half (52.6%) had practiced brushing teeth two times a day and nearly half (49.3%) reporting brushing for 2-3 minutes. Regarding use of fluoridated toothpaste, only 29.1% used fluoridated toothpaste. About 59.3% participants had decayed teeth and 3% had filled teeth. Their average knowledge scores on oral health was 7.06 (1.46) and mean DMFT score was 1.82 (1.07) (Table 1)

**Table 1.**
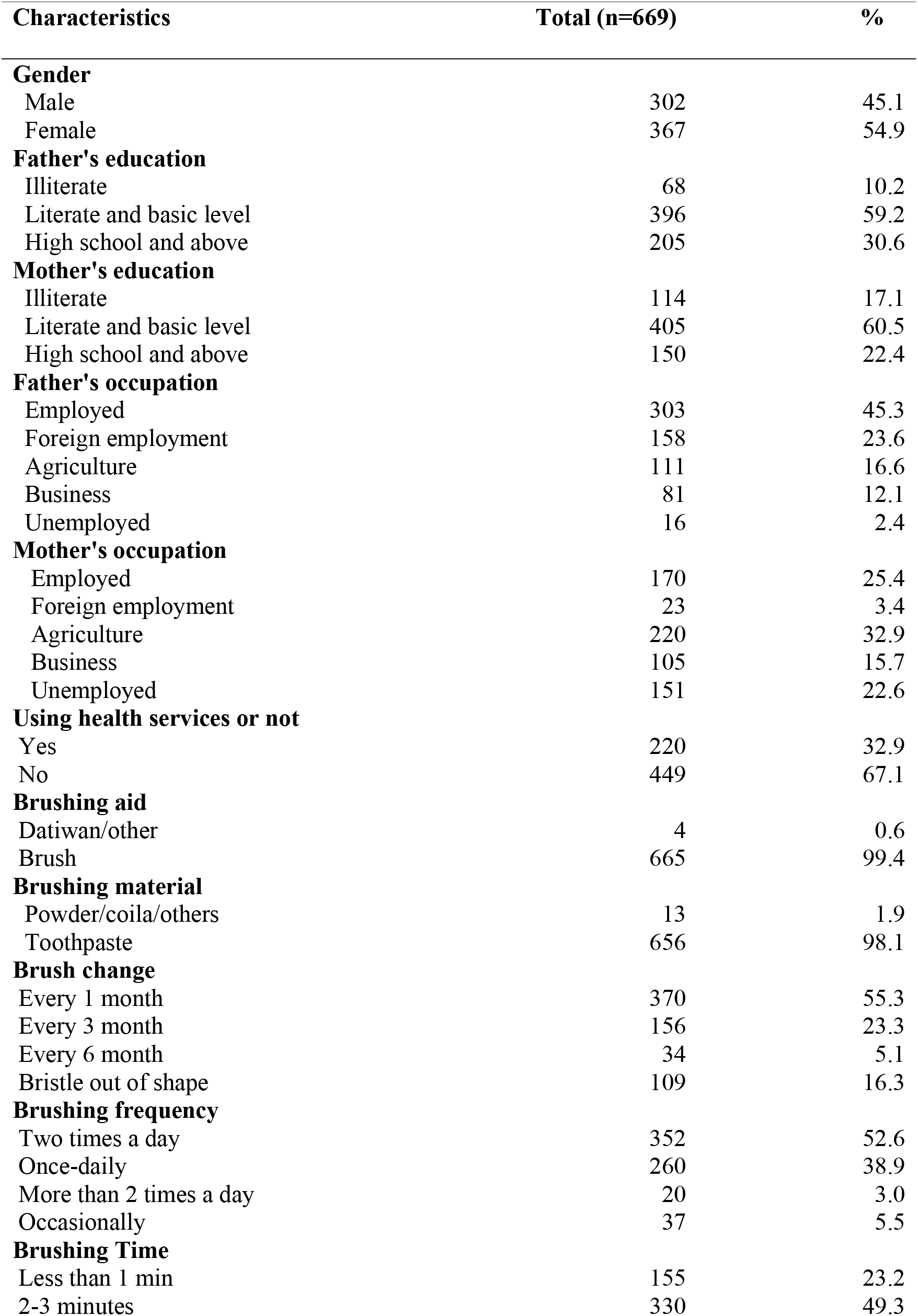
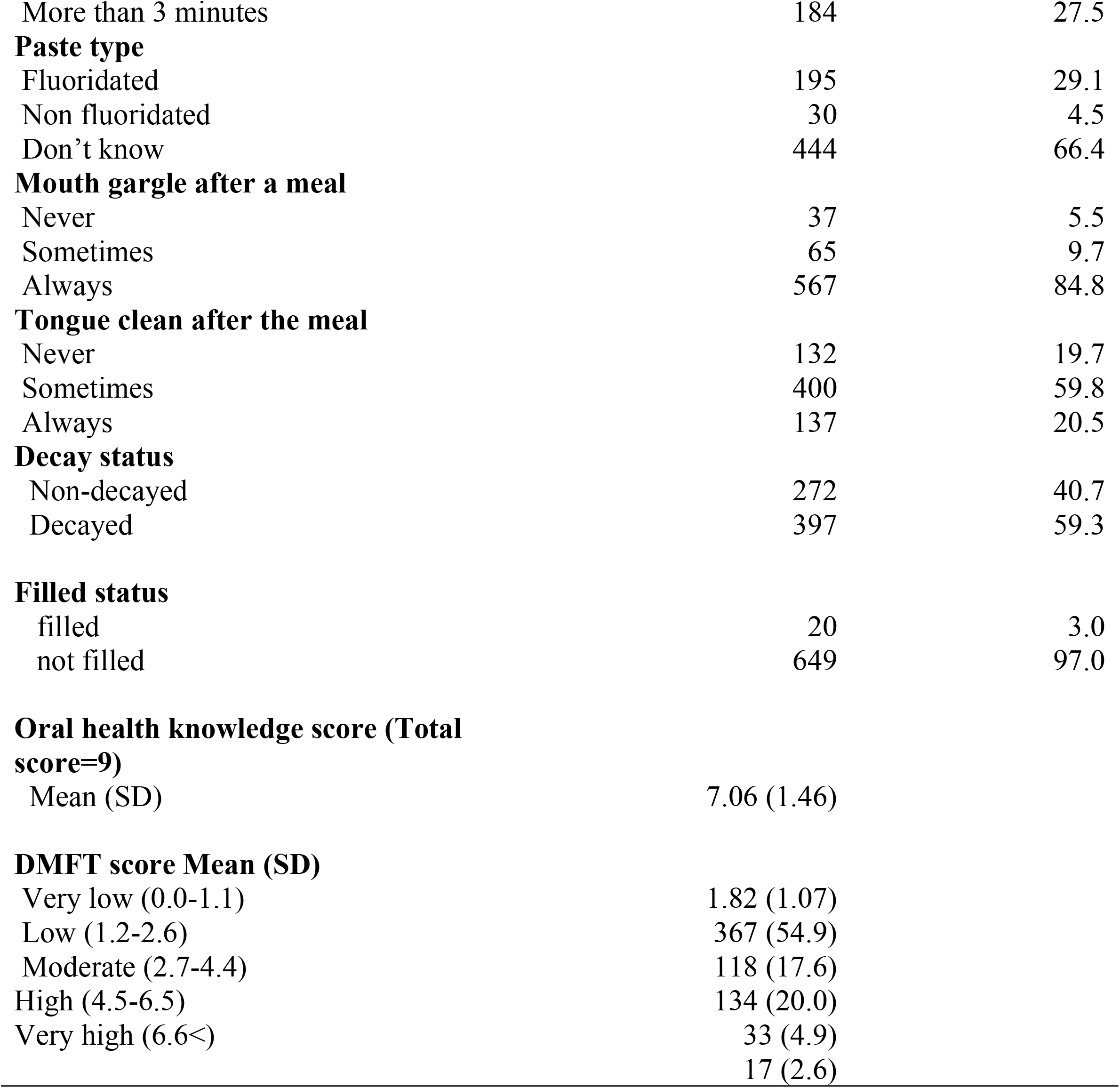
Basic characteristics of participants (n=669)

| Characteristics | Total (n=669) | % |
| --- | --- | --- |
| **Gender** |  |  |
| Male | 302 | 45.1 |
| Female | 367 | 54.9 |
| **Father's education** |  |  |
| Illiterate | 68 | 10.2 |
| Literate and basic level | 396 | 59.2 |
| High school and above | 205 | 30.6 |
| **Mother's education** |  |  |
| Illiterate | 114 | 17.1 |
| Literate and basic level | 405 | 60.5 |
| High school and above | 150 | 22.4 |
| **Father's occupation** |  |  |
| Employed | 303 | 45.3 |
| Foreign employment | 158 | 23.6 |
| Agriculture | 111 | 16.6 |
| Business | 81 | 12.1 |
| Unemployed | 16 | 2.4 |
| **Mother's occupation** |  |  |
| Employed | 170 | 25.4 |
| Foreign employment | 23 | 3.4 |
| Agriculture | 220 | 32.9 |
| Business | 105 | 15.7 |
| Unemployed | 151 | 22.6 |
| **Using health services or not** |  |  |
| Yes | 220 | 32.9 |
| No | 449 | 67.1 |
| **Brushing aid** |  |  |
| Datiwan/other | 4 | 0.6 |
| Brush | 665 | 99.4 |
| **Brushing material** |  |  |
| Powder/coila/others | 13 | 1.9 |
| Toothpaste | 656 | 98.1 |
| **Brush change** |  |  |
| Every 1 month | 370 | 55.3 |
| Every 3 month | 156 | 23.3 |
| Every 6 month | 34 | 5.1 |
| Bristle out of shape | 109 | 16.3 |
| **Brushing frequency** |  |  |
| Two times a day | 352 | 52.6 |
| Once-daily | 260 | 38.9 |
| More than 2 times a day | 20 | 3.0 |
| Occasionally | 37 | 5.5 |
| **Brushing Time** |  |  |
| Less than 1 min | 155 | 23.2 |
| 2-3 minutes | 330 | 49.3 |
| More than 3 minutes | 184 | 27.5 |
| **Paste type** |  |  |
| Fluoridated | 195 | 29.1 |
| Non fluoridated | 30 | 4.5 |
| Don't know | 444 | 66.4 |
| **Mouth gargle after a meal** |  |  |
| Never | 37 | 5.5 |
| Sometimes | 65 | 9.7 |
| Always | 567 | 84.8 |
| **Tongue clean after the meal** |  |  |
| Never | 132 | 19.7 |
| Sometimes | 400 | 59.8 |
| Always | 137 | 20.5 |
| **Decay status** |  |  |
| Non-decayed | 272 | 40.7 |
| Decayed | 397 | 59.3 |
| **Filled status** |  |  |
| filled | 20 | 3.0 |
| not filled | 649 | 97.0 |
| **Oral health knowledge score (Total score=9)** |  |  |
| Mean (SD) | 7.06 (1.46) |  |
| **DMFT score Mean (SD)** |  |  |
| Very low (0.0-1.1) | 1.82 (1.07) |  |
| Low (1.2-2.6) | 367 (54.9) |  |
| Moderate (2.7-4.4) | 118 (17.6) |  |
| High (4.5-6.5) | 134 (20.0) |  |
| Very high (6.6<) | 33 (4.9) |  |
|  | 17 (2.6) |  |

### Total Knowledge score, DMFT score and Total Decayed score by sociodemographic factors

Total DMFT score was significantly higher among females compared to males (Male: Mean=1.47 (SD=1.78) Vs female: mean=1.90 (SD=1.95); p=0.003). Similarly total decayed score was also higher among females compared to males (Male: Mean=1.24 (SD=1.53) Vs female: mean=1.62 (SD=1.75); p=0.003). Knowledge score was also higher among females compared to male (Male: Mean=6.86 (SD=1.62) Vs female: mean=7.25 (SD=1.27); p<0.001).

Those who did not visit hospitals had lower DMFT scores compared to those who visited hospitals (Hospital visit: Mean=1.99 (SD=1.99) Vs No hospital visit: mean=1.57 (SD=1.82); p=0.007). The knowledge level was lower among those who did not visit hospitals (Hospital visit: Mean=7.25 (SD=1.22) Vs No Hospital Visit: mean=6.97 (SD=1.55); p=0.018).

Knowledge level was higher among the children whose fathers had education level high school and above as compared to those whose parents were illiterate. (Father’s education high school and above: Mean=7.20 (SD=1.30) Vs illiterate: mean=6.62 (SD=1.95). (Table 2)

**Table 2.**
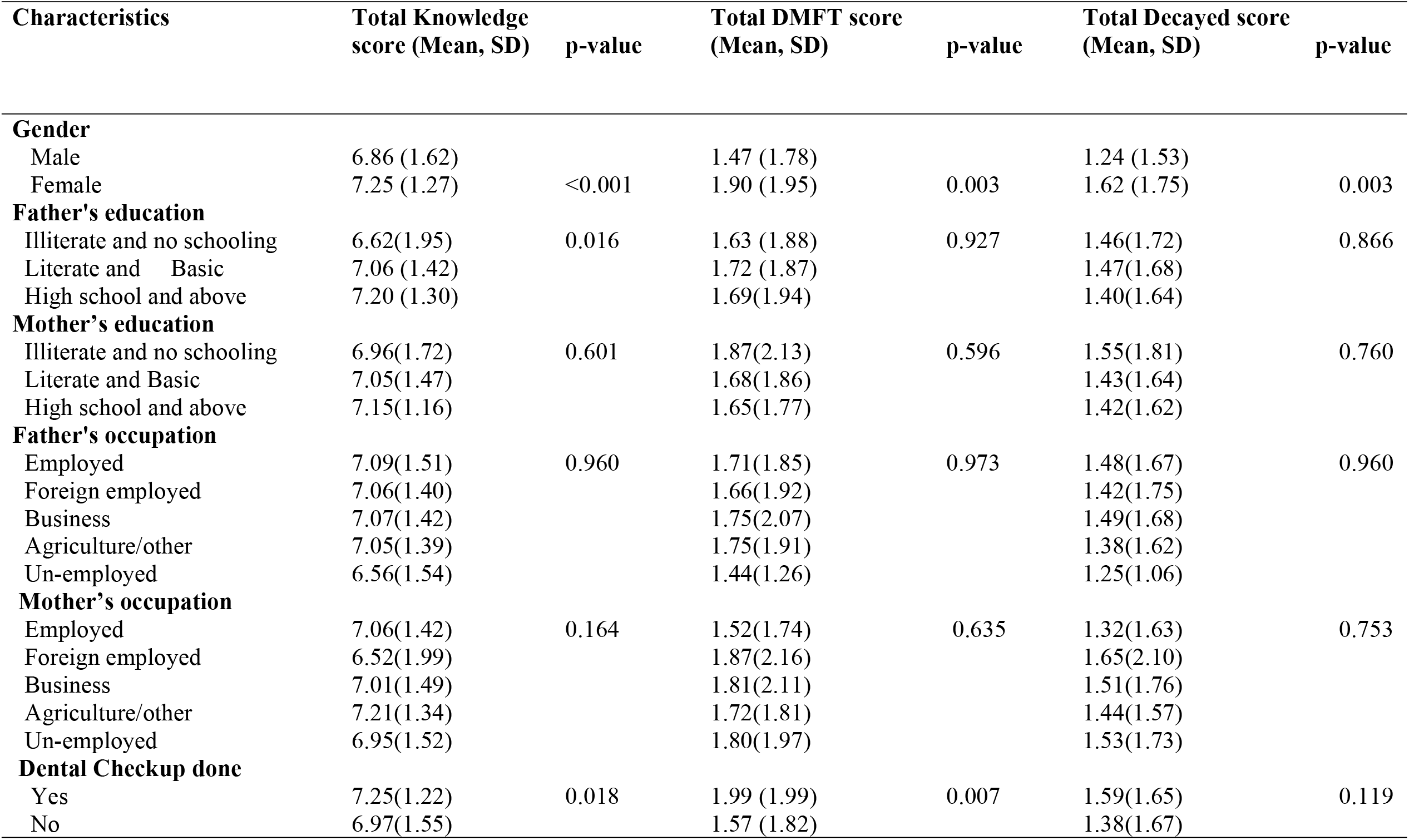
Total Knowledge score, DMFT score and Decayed score, by sociodemographic factors.

### Knowledge on oral health and DMFT score and Decayed score

Total decayed score was higher among those who did not have knowledge that fluoride prevents decay compared to those who had knowledge about it. (Knowledge about fluoride prevents decay: Mean=1.21 (SD=1.54) Vs No knowledge: mean=2.13 (SD=2.13). Those who do not have knowledge that gum diseases affect teeth had higher decayed score compared to those who had knowledge. (Knowledge about gum disease affects teeth Mean=1.37 (SD=1.59) Vs No knowledge: mean=2.30 (SD=2.02). (Table 3)

**Table 3.**
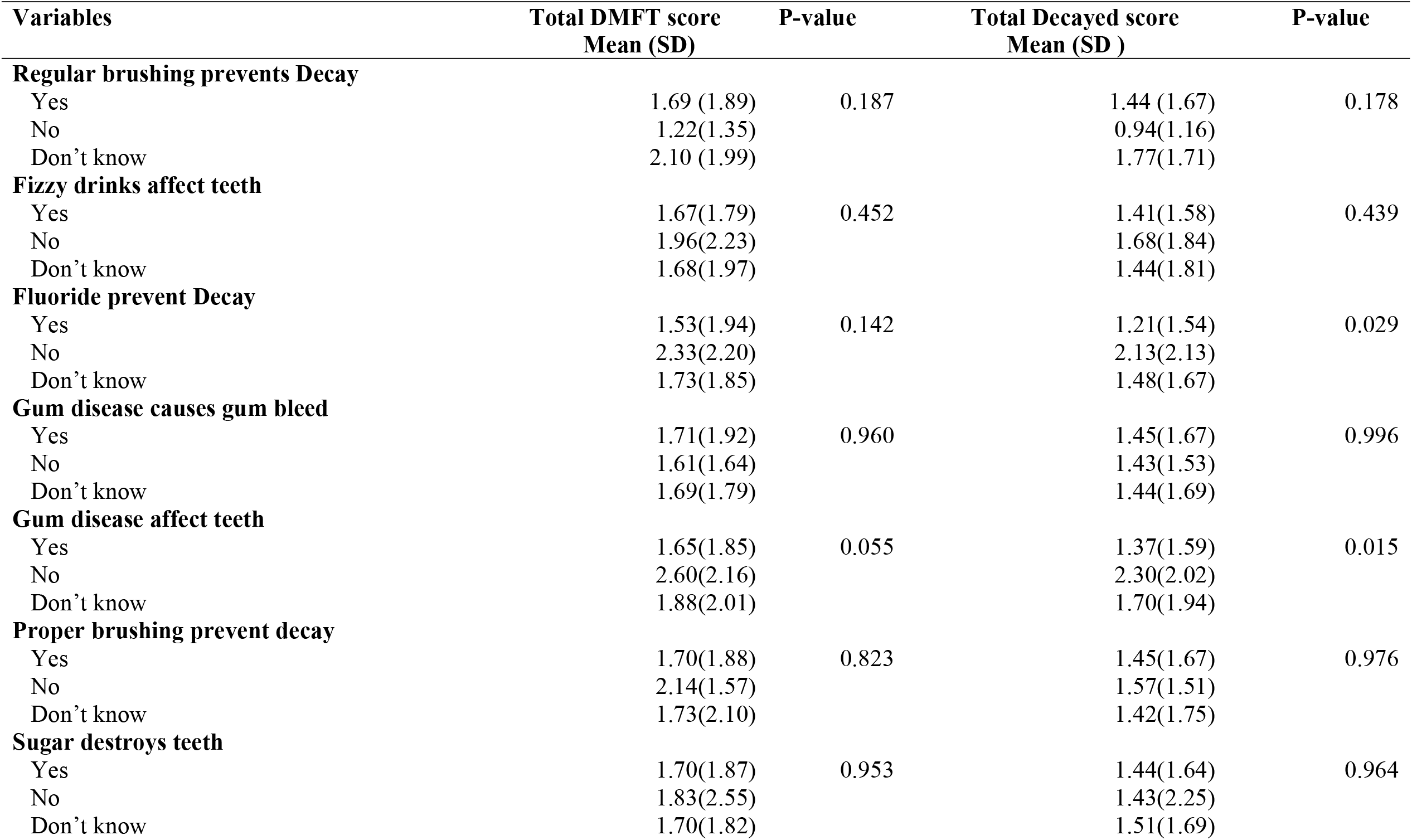
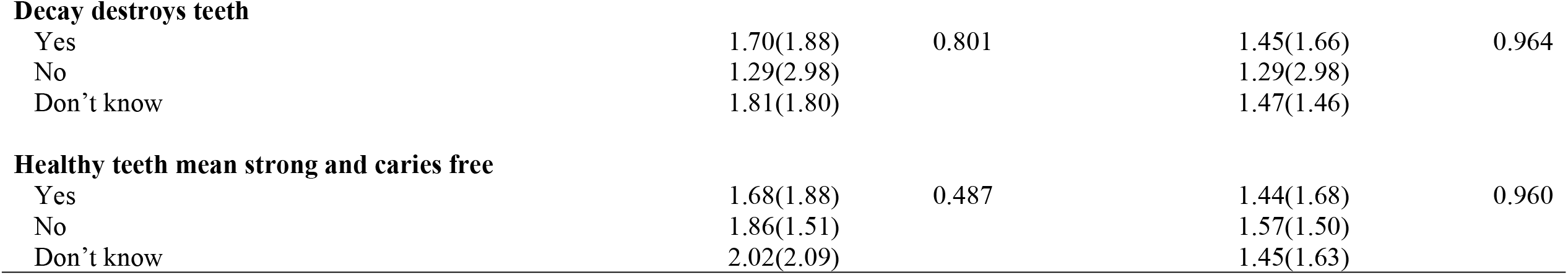
Knowledge on oral health affecting DMFT score and Decayed score (P-value by ANOVA)

#### Practice on oral health on DMFT score, Decayed score and total knowledge score (P-value by ANOVA)

Those who brushed their teeth two times a day had a higher knowledge score compared to those who brushed sometimes, once daily, two times a day, and more than two times a day. (Brushing twice a day Mean=7.22 (SD=1.34) Vs more sometimes: mean=6.76(SD=2.00). Those who used fluoridated toothpaste had higher knowledge score compared to those who did not use fluoridated toothpaste (Fluoridated toothpaste=7.58 (SD=1.32) Vs non fluoridated toothpaste: mean=7.03 (SD=1.62).

School children who had the practice of mouth gargling always had higher knowledge scores compared to those who never practiced. (Mouth gargled always =7.13 (SD=1.39) Vs Never: mean=6.27(SD=2.07); p=0.002). Those school children who cleaned their tongue always had higher knowledge scores compared to those who never cleaned their tongue (Tongue cleaned always=7.19 (SD=1.40) Vs never cleaned tongue: mean=6.57 (SD=1.53). (Table 4).

**Table 4.**
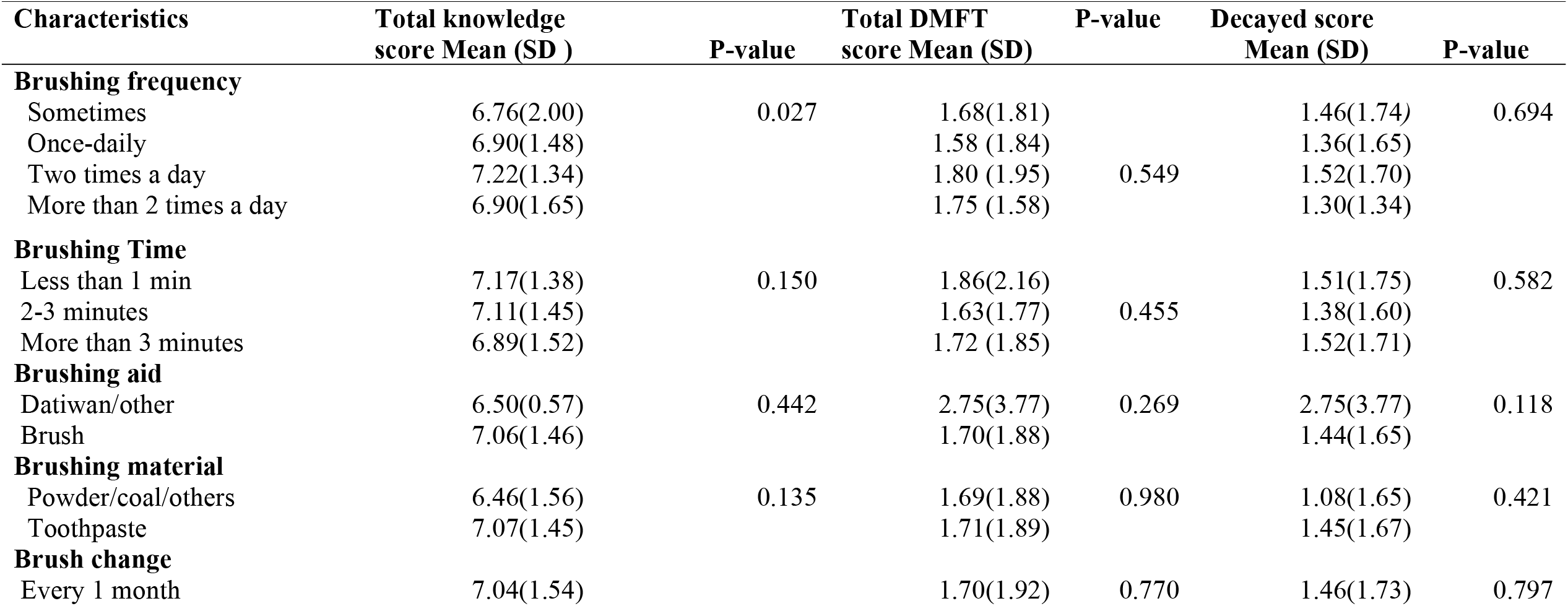
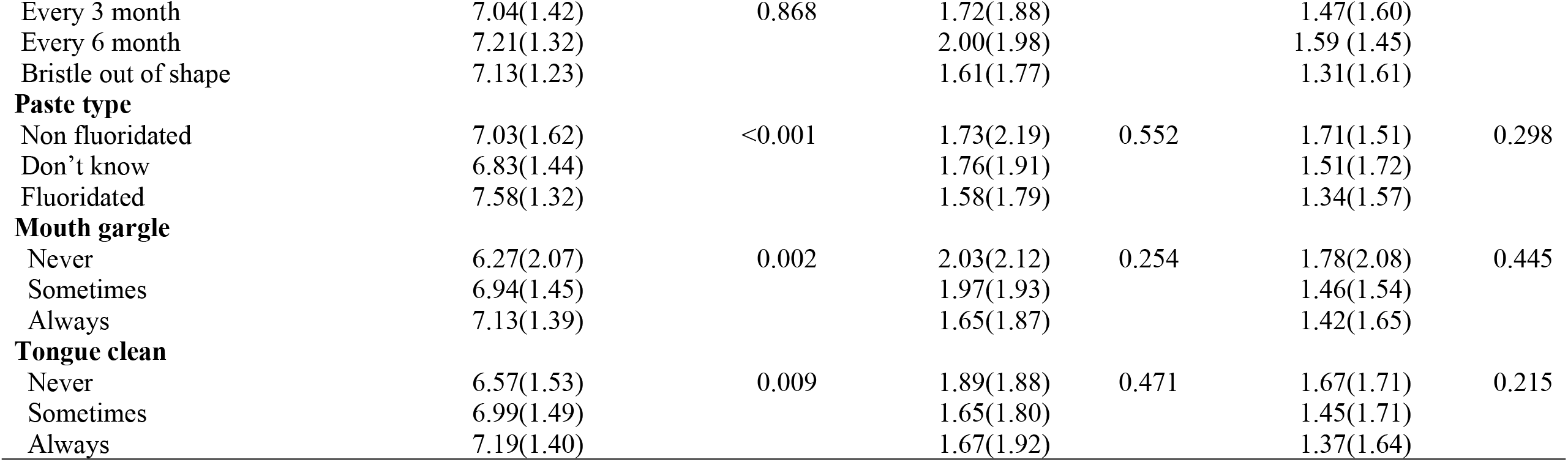
Practice on oral health on total knowledge score, DMFT score and Decayed score (P-value by ANOVA)

#### Association of socio-economic characteristics, knowledge and practices with total DMFT scores

A multiple linear regression model with gender, education level of father, education level of mother, occupation of father, occupation of mother, previous exposure to oral health education, knowledge score, availability of water, regular brushing habit, mouth gargle after meal, and tongue cleaning after meal as independent variables was developed. The independent variables that were found to be significantly associated with DMFT are gender and total knowledge score. Females are more likely to have higher DMFT score compared to male (β-coefficient=0.43, 95% CI=0.13, 0.73, P value=0.005) and increase in knowledge score is associated a with a decrease in DMFT score (β-coefficient=-0.09, 95% CI= -0.20, - 0.10, P value=0.047). (Table 5).

**Table 5.** Multiple Regression Analysis: Association of socio-economic characteristics, knowledge and practices with total DMFT.

| Characteristics | β-<br>coefficient<sup>1</sup> | 95% Confidence Interval for $\beta$ | | P-value |
| --- | --- | --- | --- | --- |
|  |  | Lower Bound | Upper Bound |  |
| Gender (female) | 0.43 | 0.13 | 0.73 | 0.005 |
| Education level of father (below secondary) | 0.08 | -0.08 | 0.24 | 0.302 |
| Education level of mother (below secondary) | -0.07 | -0.23 | 0.09 | 0.385 |
| Occupation of father (Unemployed) | -0.03 | -0.14 | 0.09 | 0.634 |
| Occupation of mother (Unemployed) | 0.06 | -0.03 | 0.16 | 0.206 |
| Previous Exposure to OHE (no) | 0.19 | -0.12 | 0.50 | 0.232 |
| Total knowledge score | -0.09 | -0.20 | -0.10 | 0.047 |
| Availability of water (no) | 0.02 | -0.70 | 0.74 | 0.962 |
| Regular brushing habit (no) | 0.12 | -0.15 | 0.39 | 0.37 |
| Mouth gargle after meal (no) | -0.22 | -0.51 | 0.08 | 0.147 |
| Tongue cleaning after meal (no) | -0.10 | -0.30 | 0.09 | 0.291 |

<sup>1</sup>Adjusted for of gender, education level of father, education level of mother, occupation of father, occupation of mother, previous exposure to oral health education, knowledge score, availability of water, regular brushing habit, mouth gargle after meal, and tongue cleaning after meal.

## Discussion

Descriptive analysis showed that the school children who visited health institutions for oral health services were found to have higher knowledge score. Moreover, their parents also had higher level of education. Children who visited health institutions for services also were found to have higher DMFT scores. School children who had knowledge about fluoride in toothpaste had lower DMFT scores compared to those who did not. Descriptive analysis also showed that the total knowledge score was higher among those who brushed properly, who used fluoridated toothpaste, and who gargled their mouth and cleaned their tongue. Children’s knowledge score on oral health was dependent on father’s education level. Parents might have given more attention to their children about oral health and sought more oral health services, thus improving their knowledge score (9). Our findings was supported by different studies conducted among school children which has shown their parents’ education level was directly related to students higher knowledge scores on oral health (10,11), In addition, our results were similar with the findings of the study conducted among school children in Nepal (12). We also found that children who visited health institution for oral health services had higher DMFT score. Similarly high decayed score was found among the school children who visited for dental services in China (13). Visiting oral health services with high DMFT score and decayed score may indicate that most of the dental visits are due discomfort and pain rather than regular visits (14).

Multivariate analysis shows that the school children who were female and those who had a lower knowledge score were more likely to have higher DMFT scores. Adolescent girls have been given low priority for health and education in children and the gender disparity remains highly prevalent in Nepali society (15). World Health Organization (WHO) has shown that adolescent girl’s health needs were higher and they had limited information and resources of health promotion activities(16). A study conducted in India also showed higher mean DMFT scores in female students compared with male students (17,18). Similar results were found in a study conducted in Qatar in 2014(19). Higher mean DMFT scores were also found in females compared to males (11). Our results were also supported by various studies conducted in South Asian countries such as India, Nepal, Bangladesh, and Sri Lanka (15). However, our results are different from the study conducted on DMFT scores among adolescents in the United Kingdom which had shown that there was no disparity among adolescents by gender in terms of their DMFT scores (20).

This study also found that decayed score was also higher among females as compared to males. A similar result was found in the study on Iranian school children (21).

Knowledge about the need of using fluoridated toothpaste was negatively associated with DMFT scores among children. Children with lower knowledge score on using fluoridated toothpaste might have limited use of fluoridated toothpaste leading to higher DMFT scores. Similar result was seen in a study conducted in Mangalore City of India (22). In addition, a study conducted in a rural village in Nepal among Chepang children had also shown that a high number of children did not have knowledge on fluoridated toothpaste and its importance to prevent dental caries (12).

Children’s proper oral hygiene practices such as proper brushing, use of fluoridated toothpaste, and gargling leads to improved oral health knowledge scores. Oral health knowledge among children had influenced improved proper brushing habit and frequency in a study conducted in China (13). In addition, this study also reported that knowledge had a role in improving oral health practices such as gargling and cleaning the tongue. Another study showed that children with a habit of proper teeth brushing frequency with fluoride containing toothpaste had a higher knowledge on oral health and lower DMFT score(23). Our study also showed that students with higher knowledge score, who gargled their mouth and cleaned the tongue had lower DMFT score. This finding is supported by a study conducted in Ethiopia in 2021(24).

This study is the first study to measure DMFT index and oral health condition and its associated factors among school children in the Gandaki Province, Nepal. The findings on associated factors in this study is based on the reported responses in the questionnaire filled by school children. A limitation may have been that this may pose some social desirability bias and school children may have reported good knowledge and practices. To minimize this bias, the researcher himself and his research assistants attended the session before filling questionnaires and briefed the school children on all questions that were to be filled out. Further, researcher himself as a dental surgeon checked up their dental condition to measure DMFT index. The second limitation was to select the community schools in urban area and peri-urban area of a district in Gandaki Province and findings may not be appropriate to generalize in other settings in Nepal. However, the socioeconomic status of the parents and settings in Nepal are somehow similar to our study settings. That is why this study can be generalized in similar settings in Nepal.

In conclusion, this study found that gender, total knowledge score and parent’s socioeconomic status were the major factors contributing to higher DMFT scores and Decayed score. To improve the oral health condition of the school children, school children should be provided with tailor made training by school teachers and regular oral health screening be conducted.

## Data Availability

All data collected are in the manuscript.

## Acknowledgements

The study team would like to acknowledge the ethical review committee and teaching faculties of University of Cyberjaya, PhD Centre Nepal and the students who participated in the schools. We also would like to acknowledge the authorities in the government schools.

## Conceptualization

Kamal Prasad Chapain, Krishna Gopal Rampal, Khem Narayan Pokhrel

## Data curation

Kamal Prasad Chapain, Kalpana Gaulee Pokhrel, Chiranjivi Adhikari, Khem Narayan Pokhrel

## Formal analysis

Kamal Prasad Chapain, Krishna Gopal Rampal, Khem Narayan Pokhrel

## Supervision

Krishna Gopal Rampal, Khem Narayan Pokhrel

## Validation

Kamal Prasad Chapain, Krishna Gopal Rampal, Khem Narayan Pokhrel

## Manuscript preparation and finalization

Kamal Prasad Chapain, Krishna Gopal Rampal, Khem Narayan Pokhrel

## Manuscript – review & editing

Kamal Prasad Chapain, Krishna Gopal Rampal, Kalpana Gaulee Pokhrel, Chiranjivi Adhikari, Deependra Hamal, Khem Narayan Pokhrel

